# White matter characterization in regions of edema surrounding meningioma brain tumor using diffusion MRI

**DOI:** 10.1101/2025.04.07.25325393

**Authors:** Sasha Hakhu, Isaac Prentiss, Jennapher Lingo VanGilder, Parvathy Hareesh, Andrew Hooyman, Edward Ofori, Jason Yalim, Justin Hines, Gabe LaFond, Leslie Baxter, Leland Hu, Yuxiang Zhou, Kurt Schilling, Scott C. Beeman

**Author notes:** Corresponding author: Scott Beeman, Ph.D.; 550 E Orange St., Tempe, AZ 85287.

## Abstract

White matter (WM) tract detection is critical in presurgical planning of tumor resection however, standard-of-care imaging techniques including T1-weighted, T2-weighted, and diffusion tensor imaging (DTI) often fail to characterize WM tracts within regions of edema. This failure arises because edema increases the isotropic diffusion component within a voxel, reducing sensitivity to the anisotropic diffusion that reflects WM integrity and directionality. More advanced diffusion modeling techniques such as free water-corrected DTI (FW-DTI), the Standard Model of Imaging (SMI), and Neurite Orientation Dispersion and Density Imaging (NODDI) address this limitation by quantifying the diffusion signals as arising from different tissue microenvironments, allowing for separation of free water, intra-neurite, and extra-neurite contributions. These techniques better preserve directionality metrics even in the presence of edema and may enhance tractography accuracy. In this study, we use multi-shell diffusion MRI data obtained from patients with meningioma brain tumors, specifically because meningiomas typically displace rather than infiltrate the surrounding WM—allowing us to isolate the effects of edema without confounding tumor invasion. We compared fractional anisotropy (FA from DTI), FW-FA (FW-DTI), P□(SMI), and orientation dispersion index (ODI from NODDI) in edematous and contralateral healthy WM regions and evaluated tractography performance across models as well. Our results show that NODDI, SMI, and FW-DTI provide improved characterization of WM within edema, yielding comparable diffusion metrics across regions and greater tract coverage compared to DTI. These improvements highlight the potential of advanced diffusion models for preoperative mapping. Future work will extend these methods to gliomas, where infiltrative tumor margins complicate WM detection, and translation into surgical navigation workflows.

## 1. Introduction

Preoperative surgical mapping of brain structures is performed to improve the accuracy of neurosurgery and reduce the risk of complications. Precise identification of white matter (WM) pathways is particularly critical because damage to these tracts can result in irreversible functional deficits, such as motor, language, or cognitive impairments. ^1,2^ However, accurate detection of WM structures in peritumoral regions remains challenging because conventional magnetic resonance imaging (MRI) techniques, such as T1-weighted (T1-w), T2-weighted (T2-w), and diffusion tensor imaging (DTI), often fail to delineate WM when present in regions of edema, a characteristic accumulation of interstitial fluid surrounding the tumor. ^3–8^ At a fundamental level, this is due to the lack of relaxation-driven signal contrast in these regions in the case of T1- and T2-weighted imaging or a failure to separate isotropic from anisotropic diffusion within a voxel in the case of DTI. In tumor-infiltrated cases, basic anatomical knowledge does not apply as tumor-induced tissue changes complicate visual assessments. To address this, we focus on a control group of meningioma patients, where WM tracts remain uncontaminated by tumor cells. Advanced diffusion modeling techniques offer enhanced microstructural insight that is particularly relevant for clinical scenarios such as pre-surgical planning, where understanding the extent of tissue disruption is essential for minimizing damage to healthy WM tracts. Additionally, besides merely improving visibility of these tracts in images, we are focused on providing an index that is specific to neurites and unconfounded by other conditions.

Advance diffusion MRI (dMRI) methods hold promise for accurate identification of WM even in the presence of edema. dMRI is sensitive to the displacement of water molecules within the constraints of the complex tissue microenvironment and, when dMRI data are rationally acquired, one can use dMRI to make quantitative inference of tissue microstructure (e.g., WM tract directionality, axon and dendrite density, and orientation complexity) through tensor calculation and/or biophysical modeling of the dMRI data. ^9–14^ DTI, which is a common and simple tensor calculation-based implementation of diffusion MRI, ^15,16^ yields the so-called mean diffusivity (MD) and fractional anisotropy (FA) metrics, of which we will principally focus on the FA metric herein. FA is scaled from 0 to 1 and is used to quantify the degree of directionality of proton displacement in the tissue. In white matter, the diffusion-driven displacement of water is typically anisotropic (greater in one direction than the others) due to the coherent orientation of white matter bundles and hence is expected to have a higher FA value. This mechanism forms the foundation of most diffusion-based study of white matter in brain. However, the utility of DTI, which is unable to compartmentalize isotropic from anisotropic diffusion components within a single voxel, can fail to characterize white matter in pathologies that involve significant edema in WM (e.g., brain tumor). In this case, edema has the effect of increasing the interstitial water volume and thus the volume fraction of isotropically diffusing protons; this necessarily reduces the total voxel anisotropic volume fraction. ^17^

Alternatively, more advanced diffusion modeling techniques exist such as the free water-corrected diffusion tensor imaging (FW-DTI), ^18^ standard model of imaging (SMI) ^19^ and neurite orientation dispersion and density imaging (NODDI). ^20^ These techniques improve WM quantification by separating the diffusion signal from distinct tissue microenvironments namely, free water (e.g., cerebrospinal fluid, interstitial fluid, edema), intra-neurite, and extra-neurite contributions. Each technique yields a directionality-relevant metric such as FA from DTI, FW-FA from FW-DTI, P□from SMI, and the orientation dispersion index (ODI) from NODDI. In particular, FW-DTI extends conventional DTI by modeling the diffusion signal as arising from two compartments— tissue and free water. The free water compartment assumes isotropic diffusion and accounts for extracellular water. By removing this isotropic contribution, FW-DTI yields corrected diffusion metrics such as free water corrected FA. SMI represents white matter diffusion as multiple Gaussian compartments— axons as zero-radius cylinders (“sticks”) in coherent fiber bundles, extra-axonal diffusion as an axially symmetric tensor, and optionally a third compartment for cerebrospinal fluid (CSF). These compartments are combined within each voxel according to a flexible fiber orientation distribution function (ODF), with consistent fractions and diffusivities across bundles differing only by orientation. NODDI models the voxel-wise diffusion signal using a three-compartment model representing free water as a sphere, intra-neurite as a stick, and extra-neurite as a tensor—and derives ODI as a measure of neurite orientation dispersion. ODI, specifically quantifies the orientation coherence of neurites in each voxel and is analogous to the DTI-derived FA value, ^21^ with the exception that the diffusion signal fraction used to calculate this value is uncontaminated by more “isotropically diffusing” proton populations like those associated with edema.

Previous studies ^22–31^ which also carried out comparative analyses using diffusion MRI and its modeling techniques in brain tumors, demonstrate the limitations of DTI in detecting WM in peritumoral regions and reported a reduction in FA values in edematous and tumoral regions. This is consistent with the fact that edema regions have an increased isotropic volume fraction and hence yield lower FA values in those regions. While these studies recognized distinguishing parameters of white matter tissue microstructure, the results of these studies must be interpreted with caution given the inclusion of heterogeneous pathological populations (e.g., glioma, brain metastases, and meningioma). For example, while both glioma and meningioma elicit peritumoral vasogenic edema, glioma displays dramatic tumor cell infiltration into neural tissue that compromises the microstructural integrity of white matter tracts and diffusion profiles of water in the tissue. ^6,7,32^ Meningioma, however, typically occurs only within the meninges, which may displace brain tissue (mass effect ^9–11^) and drive increased hydrostatic pressure and downstream vasogenic edema, but otherwise does not infiltrate or directly alter brain parenchyma. ^33^ By evaluating individuals with meningioma, it is possible to more selectively examine the effects of edema on WM tissue, minimizing the contribution of other tumor-related factors.

The goal of this study was to evaluate the performance of advanced diffusion signal modeling techniques— DTI, FW-DTI, SMI, and NODDI in characterizing WM tracts within regions of peritumoral edema in meningioma patients. Meningiomas rarely infiltrate the brain parenchyma, providing a controlled setting in which displaced but structurally intact WM tracts can be assumed. Each model yielded a directionality-relevant diffusion metric (FA, FW-FA, P□, and ODI, respectively), and tractography was performed using the primary direction vector and corresponding metric as the seeding criterion. We hypothesized that advanced diffusion modeling techniques capable of separating isotropic and anisotropic diffusion components, particularly NODDI and SMI, would provide improved WM characterization in edematous regions, compared to standard DTI. Specifically, we expected high FA, P□, and low ODI values in healthy contralateral WM, and reduced directionality in DTI i.e., lower FA in edema-laden WM. By comparing diffusion metrics and tractography across models, we aimed to determine which methods most effectively preserve WM signal in the presence of edema.

## 2. Methods

### 2.1. Participant Population

Five adults aged 62 to 77 years old bearing radiologically confirmed meningiomas were included in this study. Participant data were acquired and anonymized by the Mayo Clinic, Phoenix, Arizona and processed and analyzed at Arizona State University. All experimental procedures were approved by each institution’s Institutional Review Board. Tumor locations included: participant 1 – frontal lobe, participant 2 – frontal lobe, participant 3 – left anterior limbic lobe, participant 4 – left occipital lobe, participant 5 – left frontal lobe (Figure 1 shows a panel of these participants’ tumor locations (in a post contrast T1), edema regions (in a T2-FLAIR), and their associated FA, ODI and P_2_ maps). All meningioma cases included in this study were histopathologically confirmed primary tumors. These comprised WHO Grade I (transitional and angiomatous subtypes) — subtypes were determined through standard histological evaluation, including immunohistochemical staining (e.g., SSTR2, GFAP), following surgical resection.

**Figure 1.**
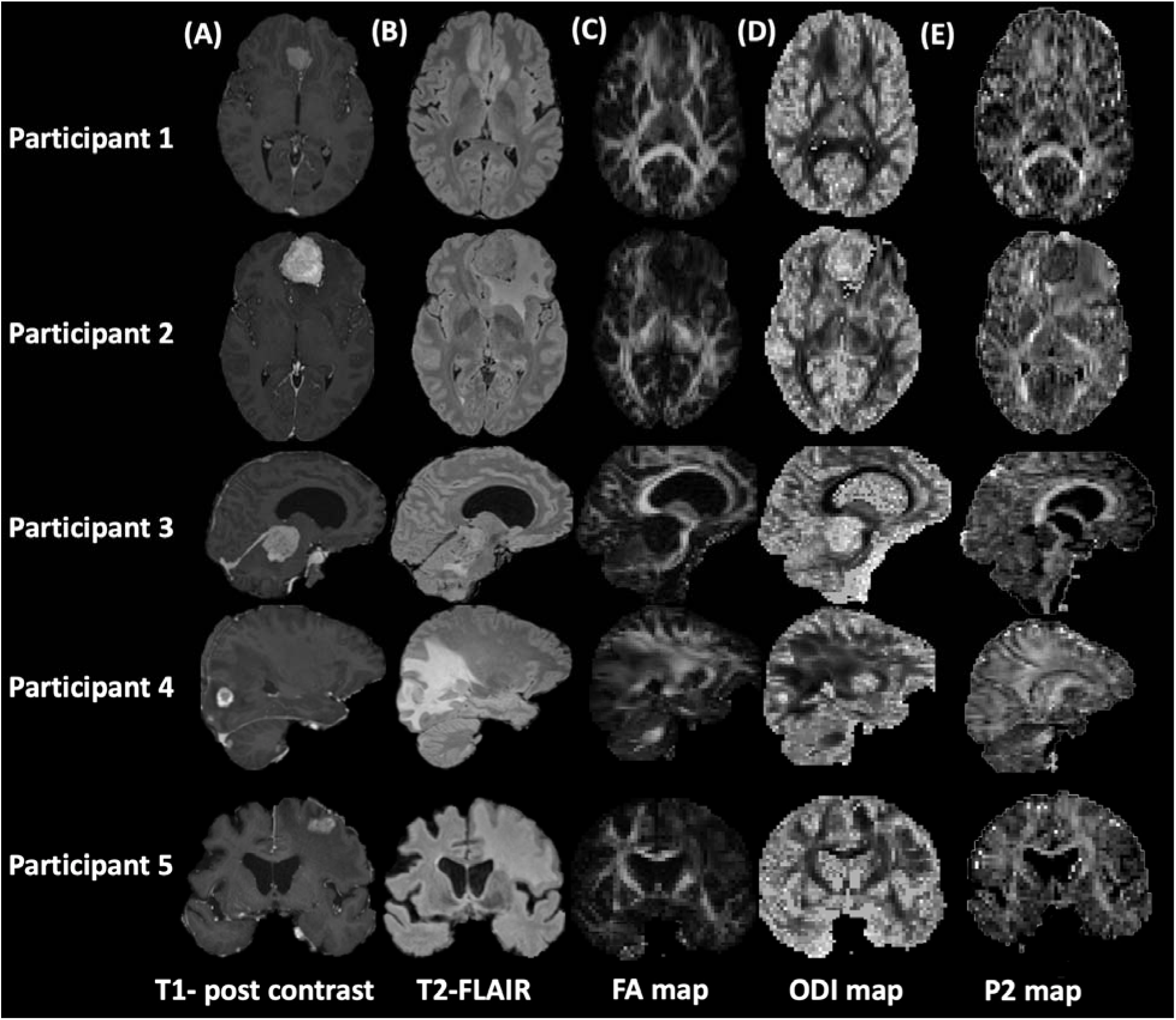
Representative (A) post-contrast T1-weighted images, (B) T2-weighted FLAIR images, (C) FA maps, (D) ODI maps and (E) P_2_ maps are shown. Post-contrast T1-weighted images best reflect tumor location and T2-weighted FLAIR images best reflect tumor plus edema location. DTI’s FA (where WM is typically represented by a brighter signal intensity) fails to robustly characterize WM tracts though regions of edema (seen as hyperintense signal traced in T2-FLAIR images) whereas NODDI’s ODI (where WM is represented by a darker signal intensity) retains WM structure irrespective of edema presence. Similarly, SMI’s p2 map (where WM is typically represented by a brighter signal intensity) succeeds. Representative image plane was chosen on a per-patient basis to best reflect the lesion.

### 2.2. Neuroimaging Data Acquisition

Imaging data were acquired on a 3T Siemens scanner using a 20-channel coil. Acquired imaging data include: (i) a three-dimensional (3D) axial T1-weighted magnetization prepared rapid acquisition gradient echo (MPRAGE) sequence, (ii) a 3D post-Gadolinium (Gd) T1-weighted MPRAGE sequence (TR/TE 1900/2.02 ms, 208 slices), (iii) a 3D T2-weighted fluid-attenuated inversion recovery (FLAIR) sequence (TR/TE 5000/389 ms, 192 slices) and (iv) multi-shell diffusion-weighted acquisitions with the following parameters: b-values (s/mm^2^) = 750 (12 directions), 1000 (18 directions), 1500 (24 directions), 2000 (30 directions) and 2500 (36 directions) (TR/TE 3664/95.6 ms, 185 slices). The total acquisition time for our multi-shell diffusion weighted imaging (DWI) protocol was 14 minutes and for the standard DTI protocol is about 10 minutes. All MR images were screened for neuropathology by a licensed neuroradiologist prior analysis.

### 2.3. Diffusion Neuroimaging Preprocessing

Only diffusion volumes with b-values of 0 and greater than 700 s/mm^2^ were included to fit the NODDI model to the data (based on its specifications) but b-values= 0, 1000 s/mm^2^ were used in the case of DTI. Data were corrected for noise (MRtrix), ^34,35^ field map distortion (Synb0) ^36^ and eddy currents (MRtrix and FSL). ^37^ Specifically, for noise correction, we used MRtrix’s *dwidenoise* tool, which applies a data-driven approach to reduce noise based on a local estimate of the data’s noise characteristics. This tool is based on the MPPCA ^35^ method. We applied Synb0-DISCO for susceptibility distortion correction, generating an undistorted b0 image that was used with *topup* to correct for geometric distortions in diffusion-weighted images caused by susceptibility effects. Additionally, *eddy* was applied to correct for eddy current distortions, motion artifacts, and slice-to-volume misalignments, while also performing outlier detection to identify slices affected by subject movement. DTI (using FSL’s DTIFIT toolbox), ^38^ FW-DTI, ^18^ SMI ^39^ and Watson-NODDI (using the NODDI MATLAB toolbox) ^40^ models were applied to preprocessed data to generate diffusion parameter maps, with FA, FW-FA, P□, and ODI maps representing directionality-relevant metrics from each method, respectively. These maps are scaled from zero to one; where high FA, high P_2_ and low ODI reflect coherent directionality of WM tracts. All volumes were then co-registered to the pre-contrast T1-weighted volume.

### 2.4. ROI Segmentation and Processing

For each participant, the tumor region of interest (ROI) was identified and manually segmented using the post-contrast T1-weighted volume. The total edema ROI (including tumor and peritumoral edema) was identified and segmented based on the hyperintense signal of the T2-FLAIR. The extra-tumoral edema ROI was derived by subtracting the tumor segment from the total edema segment. As a control, healthy-appearing contralateral white matter tracts were identified by first flipping the extra-tumoral edema ROI over the midline and then manually adjusting the outline to include only white matter containing voxels as defined on the pre-contrast T1-weighted image. These ROIs were then overlaid on diffusion maps (FA, P_2_, ODI) and the location of each ROI was confirmed by a licensed neuroradiologist (see edema appearance in these image volumes in in Figure 2). Additionally, to address the bias caused by CSF contamination in the ROI analysis, we implemented a tissue-weighted mean approach. ^41,42^ Traditional methods average NODDI metrics like NDI and ODI across all voxels, which can lead to inaccuracies in areas with partial CSF volume. Here, we use tissue fraction (TF) maps, calculated as one minus the free water fraction (FWF), to adjust for this. By multiplying NDI and ODI by the TF, extracting their weighted means, and normalizing by the mean TF, we obtain more accurate estimates of the microstructure, effectively correcting for CSF contamination. This tissue-fraction correction is similarly intrinsically accounted for in SMI-derived metrics to mitigate the influence of free water and ensure more tissue-specific estimates.

**Figure 2.**
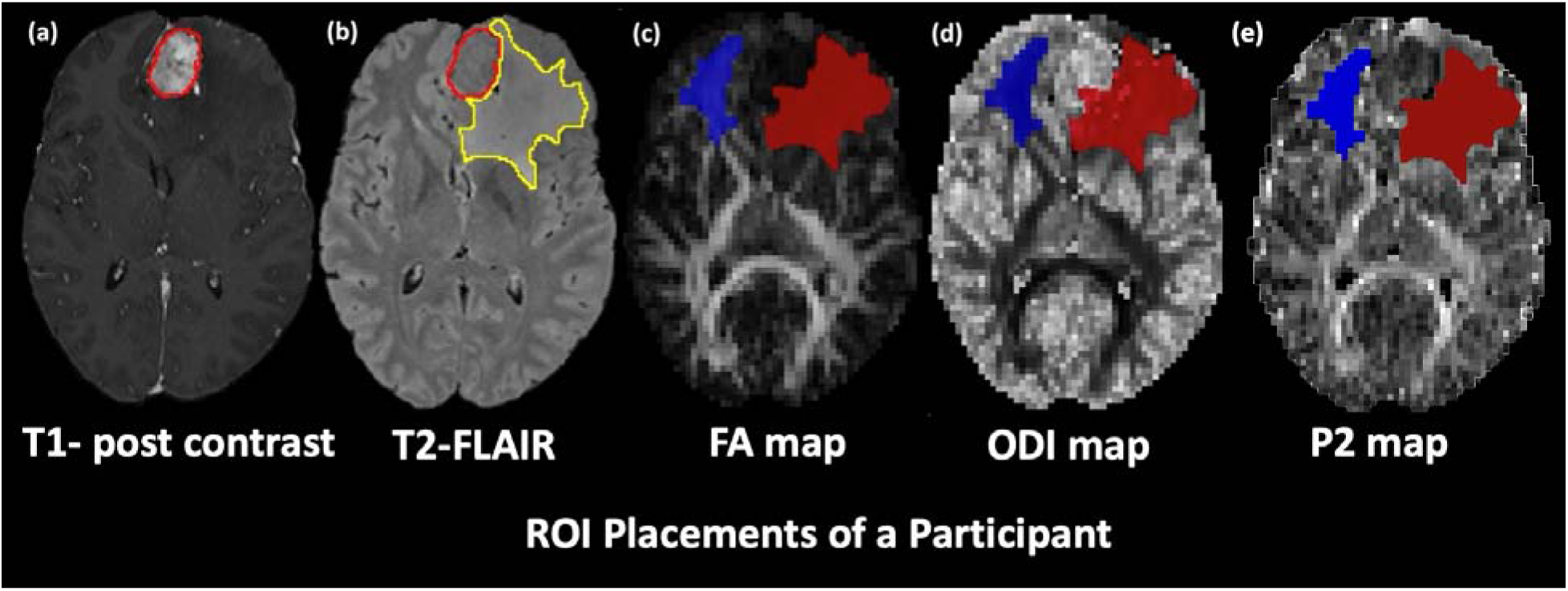
Example ROI placements on T1-post Gd and T2-FLAIR images and FA and ODI maps. (a) T1-post Gd image with tumor contour (red), (b) T2-FLAIR image with tumor (red) and edema (yellow) contours, (c) FA, (d) ODI maps and (e) P_2_ maps with edema (in red) and the contralateral region (in blue) highlighted. For each participant, the tumor contour was identified and segmented using the T1-weighed post-contrast volume and was then overlaid the T2-weighted FLAIR image; then the peritumoral edematous contour was identified and segmented based on the hyperintense signal in the T2-FLAIR. The region highlighting extra-tumoral edema was then derived by subtracting the two segmentations (i.e., FLAIR region of interest minus the post-contrast T1-weighted region of interest). As a control, healthy-appearing contralateral white matter tracts were identified by first flipping the edema ROI over the midline and then manually adjusting the outline to include only white matter containing voxels. A T1-weighted image was used to verify locations of contralateral WM tract ROIs. These ROIs were then overlaid on the diffusion maps and the location of each ROI was confirmed by a licensed neuroradiologist. Participants’ images were then co-registered to their T1-weighted image.

Voxel values were extracted from edematous and contralateral ROIs using *fslstats.* Distributions were assessed using R version 4.2.2. ^43^ A Shapiro-Wilks test was used to test the metric distributions for normality. t-tests were performed between WM metrics in edematous and contralateral non-edematous regions.

### 2.5. Tractography

For tractography, we implemented “Fiber Assigned by Continuous Tracking” (FACT) (using MRtrix’s *tckgen* package ^44^ and visualized using MI-Brain ^45^). Here, the primary direction vector was used as the input image, along with the directionality-specific metrics (DTI-FA, FW-DTI-FA, NODDI-ODI and SMI-P_2_) as seed images. Specifically, for NODDI-based tractography, the orientation dispersion index (ODI) map was subtracted from one to align its contrast with that of FA, FW-FA, and P□ maps. This step was necessary to satisfy thresholding assumptions inherent to the FACT algorithm, which interprets higher seed map values as indicating greater directionality and coherence for streamline propagation. The directionality maps, along with their respective primary direction vectors, were then used as seed images and input images, respectively. A seed initiation threshold of 0.88 and a tract termination cutoff of 0.84 were applied across all directionality maps (FA, FW-FA, P□, and 1–ODI). These values were selected based on a previously reported correspondence between FA and ODI, ^46^ where an FA of 0.50 approximates an ODI of 0.16, and an FA of 0.60 approximates an ODI of 0.12. Subtracting these ODI values from one yields corresponding thresholds of 0.84 and 0.88, ensuring consistency in directionality interpretation across models. ^46^ Ventricle and white matter masks were created (using FSL) to exclude any streamlines in the region of ventricles and to restrict streamlines to the bounds of the white matter. A total of 10,000 streamlines were generated for all methods used here.

To assess fiber tracking through edematous tissue, we calculated the mean edema tract fraction for each participant, defined as the proportion of voxels within the edema ROI that contained at least one streamline. ^47^ This metric reflects the spatial coverage of tractography in edema, with lower values indicating reduced tracking. Using MRtrix3, *tckmap* generated a streamline density map, which was thresholded (*mrthreshold*) to retain voxels with ≥1 streamline. This binary map was masked to the edema ROI (*mrcalc*), and the number of streamline-containing voxels (*mrstats*) was divided by the total edema voxel count to yield the tract fraction.

## Results

Quantitative differences between edematous and contralateral regions were assessed using FA, ODI, and P2 metrics derived from DTI, NODDI, and SMI models, respectively. Percentage differences were computed for each ROI using a standardized formula (Equation 1), and the results are summarized in Table 1. Additionally, tractography-derived edema tract fractions, calculated using model-specific directionality metrics (DTI-FA, FW-DTI-FA, NODDI-ODI, and SMI-P2), are reported in Table 2 across all participants.

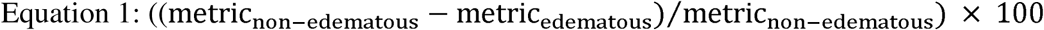

**Table 1.**
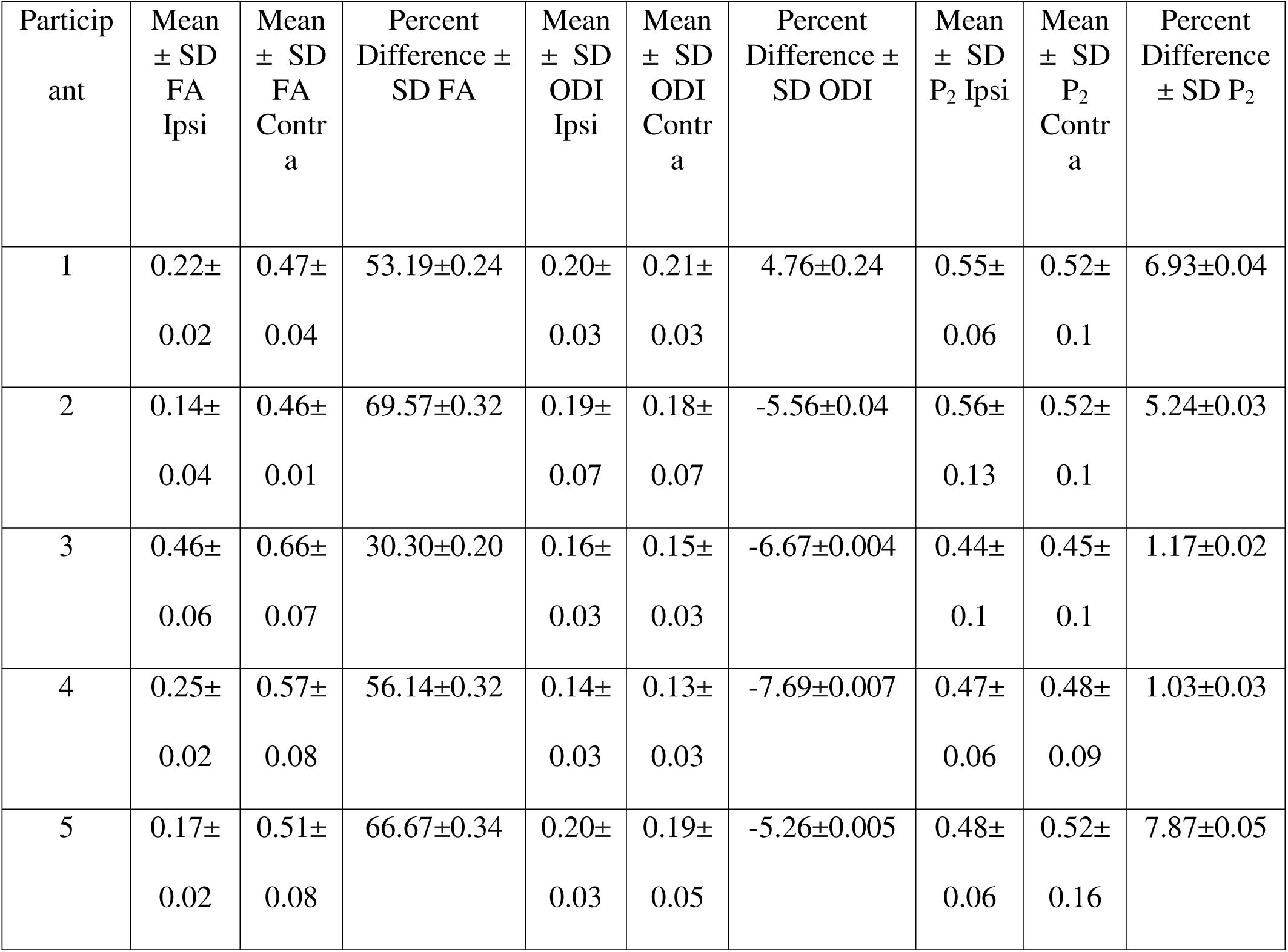

**Table 2.**

| Participant | Mean edema tract fraction FA | Mean edema tract fraction FW-FA | Mean edema tract fraction ODI | Mean edema tract fraction P<sub>2</sub> |
| --- | --- | --- | --- | --- |
| 1 | 0.57 | 0.95 | 0.94 | 0.95 |
| 2 | 0.59 | 0.93 | 0.96 | 0.91 |
| 3 | 0.39 | 0.89 | 0.95 | 0.77 |
| 4 | 0.51 | 0.86 | 0.85 | 0.74 |
| 5 | 0.29 | 0.78 | 0.94 | 0.70 |

While FA and P_2_ values were high in contralateral white matter tracts and low ODI (as expected due to its inverse scaling as compared to FA and P_2_), as shown in Figure 3, FA was lower in edematous white matter tracts whereas the P_2_ and ODI values remained approximately in the same range for both regions. There were no significant differences in the ODI or P_2_ values between edematous and non-edematous regions (β = 0.0007, 95% CI = [-0.03; 0.03], p = 0.96) but differences were significant in the FA values within these regions (β = 0.29, 95% CI = [0.16; 0.42], p = 0.0008). This lack of significant difference in P□ and ODI across hemispheres is a desirable outcome, as it reflects the ability of SMI and NODDI to maintain stable directionality metrics even in the presence of edema.

**Figure 3.**
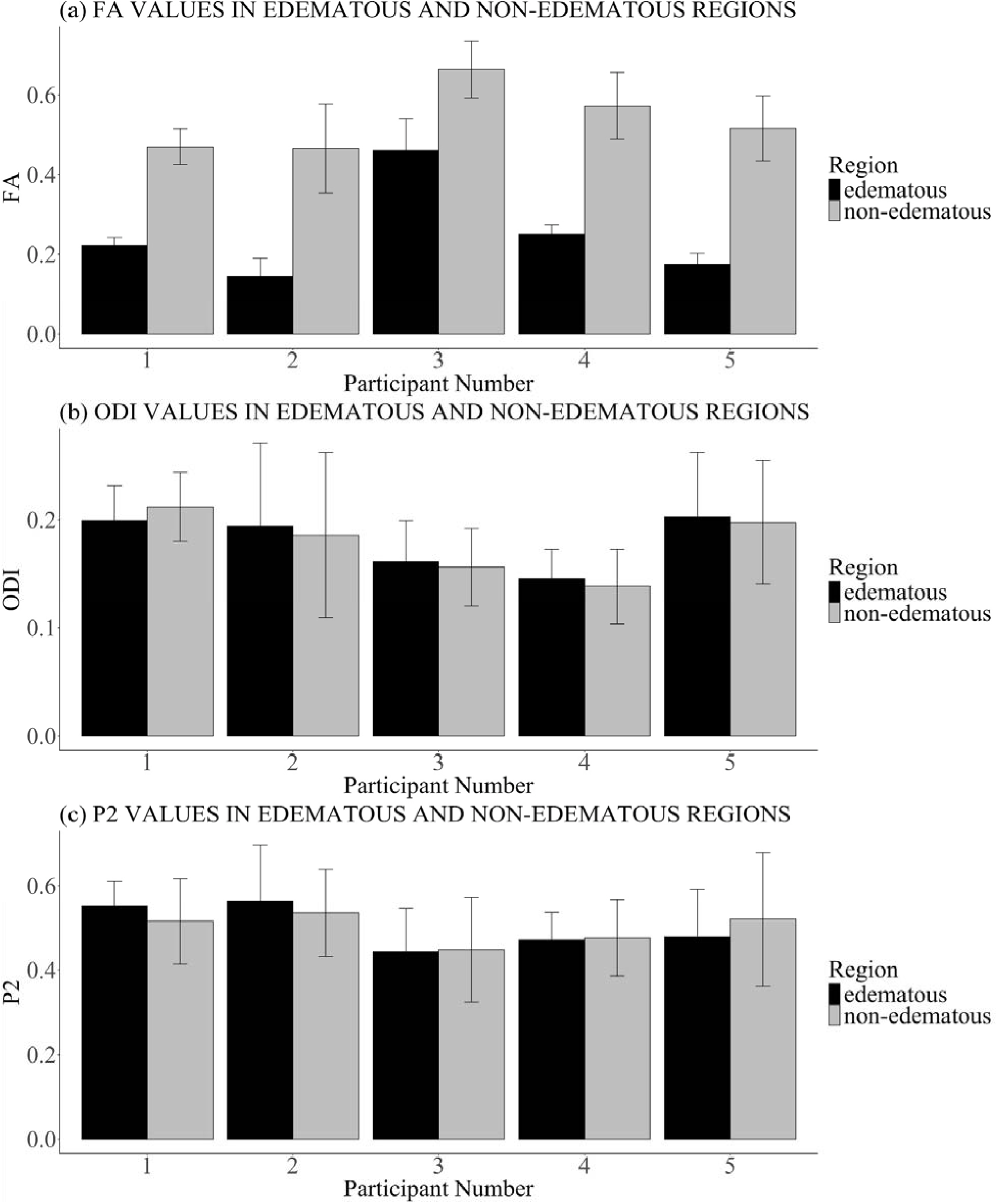
Values between white matter regions of interest in edematous and non-edematous regions for (a) DTI-FA maps (b) NODDI-ODI maps (c) SMI-P_2_ maps.

To further characterize microstructural changes in edematous white matter, we examined NODDI-derived metrics alongside volumetric analyses. Supplementary material includes Figure S1 (a) NODDI-derived isotropic volume fraction (fISO) map and Figure S1 (b) which shows the neurite density index (NDI) map for a participant with left hemisphere edema. The accompanying plots illustrate average voxel-wise fISO and NDI values in edematous regions compared to the contralateral healthy side. fISO values appear to capture intra-voxel levels of tumor-induced edema. This is supported by lower NDI values in edematous areas, which reflects an approximate inverse relationship between neurite density and edema volume. Further supporting this, volumetric analyses using T1-weighted images revealed that the edematous regions were consistently larger than their contralateral counterparts. On average, edematous regions showed a mean volume of 1067 ± 571 mm³, compared to 791 ± 472 mm³ on the contralateral side. This corresponds to an approximate mean increase of 39.7% (± 5%) in volume on the edematous side. This significant increase suggests the spread of interstitial fluid into white matter (WM), leading to volumetric expansion of the tissue.

Tractography results showed clear differences across models in their ability to track fibers through edematous tissue. DTI-FA failed to consistently generate tracts within edema, while FW-DTI, SMI and NODDI-based methods were able to produce streamlines in these regions. Figure 4 shows representative tractography results, highlighting the contrast between DTI the other advanced diffusion modeling-based tractography methods successfully characterized tracts within the edematous regions and, Figure 5 summarizes the edema streamline volume fraction across participants for each model, and

**Figure 4.**
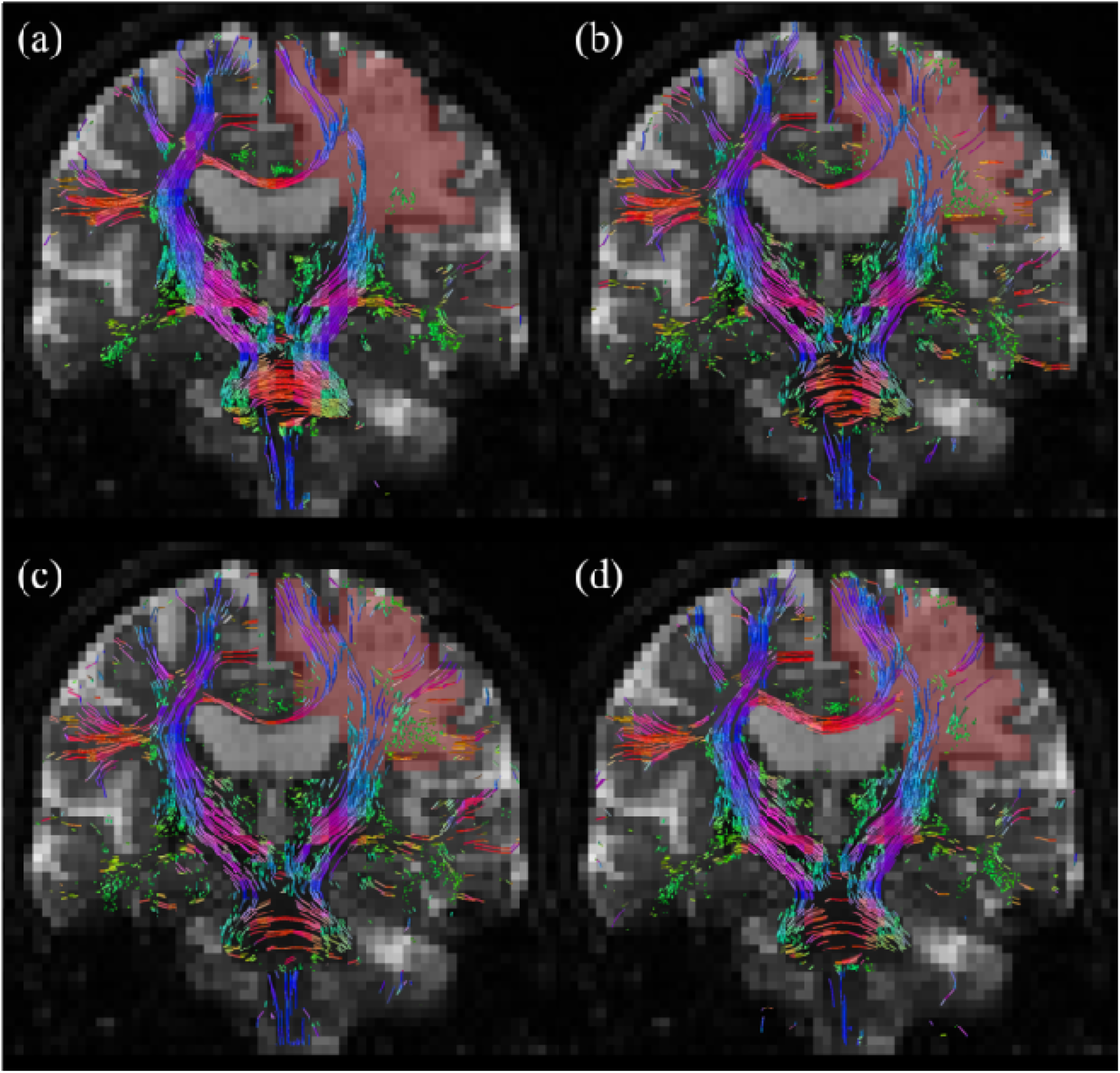
Comparison of whole brain tractography viewed along a coronal slice between using the deterministic algorithm with (a) Standard DTI, (b) FW-DTI (c) NODDI-based (d) SMI-based methods for fiber tracking through region of edema along a coronal slice.

**Figure 5.**
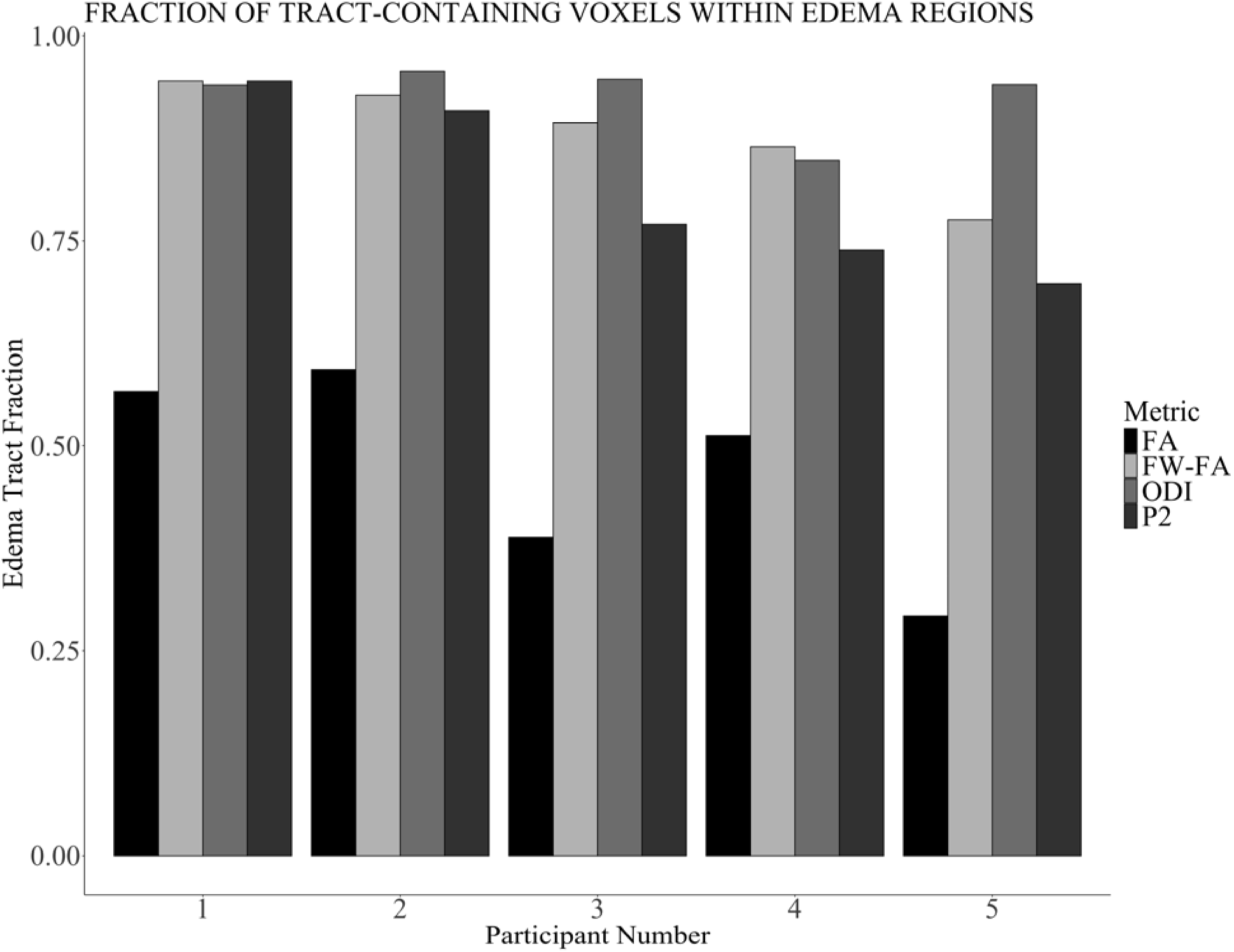
Streamline volume fraction comparison for participants across deterministic tractography algorithms implemented using— DTI, FW-DTI, NODDI and, SMI in regions of edema across all participants.

## 3. Discussion

In this study, we focused on using advanced diffusion modeling techniques to characterize WM in a controlled trial containing only meningioma cases where no WM disruption or tumor invasion is expected in regions of edema. ^17,20,33,48^ This was supported by radiological evaluation, including independent review and confirmation by a certified neuroradiologist, who verified that the regions of edema did not overlap with tumor margins or areas of suspected WM compromise. This approach contrasts with previous studies that have compared advanced modeling techniques and DTI across heterogeneous populations involving various tumor types, where the surrounding tissue environment can be more complex (often involving destruction of WM tracts). ^23,25,26,28^

Consistent with our hypothesis, advanced diffusion modeling techniques—FW-DTI, NODDI, and SMI, provided improved white matter characterization in edematous regions (please refer to Table 1 and Figure 3). This improvement comes from these methods’ ability to explicitly separate isotropic and anisotropic diffusion components within a voxel. Directionality metrics reflected this distinction i.e., DTI-derived FA showed large percent differences between edematous and contralateral WM (∼55%, range: 30–70%), indicating high sensitivity to edema. In contrast, NODDI-derived ODI and SMI-derived P□showed minimal differences (<7%), suggesting high sensitivity to WM and greater robustness to free water effects. Such stability suggests that these models are less susceptible to isotropic contamination and may provide a more reliable representation of underlying white matter microstructure in pathological contexts. These enhanced diffusion representations translated into improved tractography performance (please refer to Table 2 and Figure 5). While DTI-based streamlines were sparse and fragmented in edematous areas (mean tract volume fraction = 0.47), FW-DTI, NODDI, and SMI produced more continuous and extensive tracts (volume fractions = 0.88, 0.93, and 0.81, respectively). Together, these results support the value of advanced modeling for preserving white matter signal in regions affected by edema.

While significant differences in diffusion metrics between edematous and non-edematous white matter likely reflect sensitivity to edema-related factors (e.g., increased free water), these differences do not always equate to precise characterization of white matter microstructure. Such findings may instead highlight the confounding influence of edema on diffusion measures, emphasizing the need for cautious interpretation in clinical contexts. Although white matter properties are generally expected to be comparable across hemispheres, statistical significance alone may be influenced by sample size and does not imply statistical similarity. Our objective is not to claim statistical equivalence but to demonstrate that advanced diffusion models yield more consistent and reliable estimates of white matter microstructure amidst edema. Conversely, reduced fractional anisotropy (FA) values in standard DTI likely reflect its known limitations in distinguishing isotropic from anisotropic diffusion within a voxel. Importantly, this does not diminish the value of FA as a sensitive metric but underscores its limitations in accurately representing tract integrity in edematous tissue.

Additionally, while mean tract volume fraction was used to compare tractography performance across models, we acknowledge that it is a surrogate metric and does not confirm anatomical accuracy. The gold standard remains validation of the underlying diffusion maps against histological reference. Also, free water-corrected FA maps often exhibit zero-valued voxels in regions of peritumoral edema due to removal of the isotropic free water compartment. ^18^ These zero-value voxels reflect the absence of tissue-specific anisotropy. However, the tracking algorithm ^49^ utilizes both a seed image and a primary direction vector as the input image. In FW-DTI tractography, although the FW-FA map serves as the seed mask, the primary eigenvector derived from the FW-corrected tensor guides streamline propagation. This means that even if a region lacks FA magnitude, preserved directional coherence in adjacent voxels can still support tracking through or around the edema. Therefore, the success of FW-DTI tractography lies not in the seeding density within edema, but in the retention of coherent directional information across the voxel field. Ongoing work includes methods such as CSD (constrained spherical deconvolution). ^50^

In the context of threshold-driven tractography, the study by Chong et al. (2021), ^22^ for example, highlighted the use of NODDI to enhance tractography in regions affected by peritumoral edema, demonstrating that NODDI-based tractography could visualize fiber tracts not detectable by standard DTI. They calibrated an optimal ODI threshold using the deterministic fiber tracking algorithm for the corticospinal tract (CST) in healthy subjects to then assess the differences between DTI and NODDI-derived tractography in a population of brain tumor subjects (a mixed population of high- and low-grade tumors like gliomas and meningiomas). In contrast to tumor-infiltrated regions, where tumor cells complicate assessments, our study specifically examines WM regions in subjects with non-invading meningioma tumors wherein WM is unaffected by tumor cells; for this reason, we are able to isolate the effects of edema from other likely confounds like tumor infiltration and demyelination. By incorporating these controls and systematically comparing tractography outcomes across DTI, FW-DTI, SMI and NODDI, we demonstrate the superior performance of advanced diffusion modeling methods in viewing tract integrity in the presence of edema without any other uncertainties associated with invasive tumors.

Our study focused on examining white matter microstructure in regions affected by peritumoral edema rather than on specific neuroanatomical tracts. To achieve this, we used manually segmented ROIs to cover relevant regions masking edema. While a more comprehensive reconstruction of white matter tracts could provide additional insights, our primary objective was to compare diffusion models in the context of edema-related microstructural changes. Therefore here, anatomical specificity was not the central focus, but rather the broader characterization of white matter within edematous regions. For instance, while participant 3 had a meningioma seemingly affecting deeper structures, the presence of asymmetrical edema was particularly relevant, as it allowed us to assess white matter changes and compare it to contralateral unaffected regions. Additionally, some inter-subject variability was observed in contralateral WM metrics, potentially reflecting anatomical differences or minor partial volume effects. While such factors are important to consider when interpreting WM measurements, particularly in clinical research settings, they do not detract from the overall trends observed across participants in our analyses.

The DTI protocol used in our study reflects standard clinical acquisition settings, ensuring broader applicability of our findings. While higher b-values (e.g., 1500 s/mm²) can enhance microstructural characterization, they also introduce lower SNR and increased motion sensitivity, which can be limiting in clinical settings. ^51,52^ Similarly, the 18 diffusion directions used for DTI reflect standard clinical practice, balancing acquisition time and data quality for broader applicability. ^53^ NODDI, for example, requires more directions to accurately model microstructure, especially in regions affected by pathology. This is due to the different complexity of the two methods. Here, the focus was on comparing different modeling methods under typical acquisition constraints rather than optimizing DTI for improved performance. The key difference being that multi-shell diffusion data allows for models like SMI and NODDI to provide more reliable information about tissue microstructure, even in areas like edema.

Historic challenges to deploying these methods include collection of multi-shell datasets which required increased acquisition times; however, the emergence of multiband, ^54^ parallel imaging, ^55^ and compressed sensing ^56^ acquisition schemes has dramatically reduced acquisition times to the point where dMRI datasets that satisfy more advanced diffusion modeling techniques can now be acquired in approximately five minutes. ^30^ Importantly, multi-shell datasets open the door to other models but don’t preclude DTI and its use in standard clinical procedures. While in this work we have done our best to ensure the presence of *only* edema in imaged tissue via selective inclusion of only meningioma-bearing participants, future studies will need to focus on the more complex cases of invasive tumors like high grade gliomas. The study of invasive primary brain tumors in this context would also greatly benefit from complementary rodent studies in which dMRI results can be directly validated against histologic and assay studies; studies which are ongoing in this laboratory. Finally, future work will attempt to involve expansion into other biophysical models of diffusion in brain tissue which have great data demands, including variations on the Standard Model ^19^ like the Soma And Neurite Density Imaging (SANDI) model ^57^ and the Neurite Exchange Imaging (NEXI) ^58^ in both clinical and preclinical settings.

## 5. Conclusions

In this study, we compared the performance of standard DTI with advanced diffusion modeling techniques i.e., FW-DTI, SMI, and NODDI for characterizing white matter in peritumoral edema. Directionality metrics and tractography derived from these advanced methods demonstrated improved detection and mapping of displaced but intact WM tracts, as evidenced by lower percentage differences (between edema-displaced WM and contralateral intact WM), higher spatial coverage and more coherent streamlines through edematous regions. This improvement stems from their ability to separate isotropic (e.g., edema-related) and anisotropic (WM-related) diffusion components within a voxel thus overcoming a key limitation of standard DTI. These findings highlight the potential of biophysical diffusion models, particularly NODDI and SMI, to enhance presurgical planning in cases where edema obscures WM, and motivate future applications in glioma, where differentiating infiltrative tumor from edema is even more clinically critical.

## Data Availability

All data produced in the present study may/may not be available upon reasonable request to the authors.

## Acknowledgments

This work was supported by NIH Grant R37CA288396 (S.C.B.); NIH grant U01CA220378 (L.S.H); NIH grant U01CA250481 (L.S.H.); NIH grant R01CA264992 (L.S.H.); NIH grant R01CA221938 (L.S.H.); NIH grant R21NS082609 (L.S.H.); K01 EB032898 (K.S.); the Mayo Clinic & ASU Alliance for Health Care— Collaborative Research Seed Grant Program; the ASU-Mayo Summer Residency program (SCB, LSH) and the Arizona Biomedical Research Center (S.C.B.).

---

**Figure S1.**
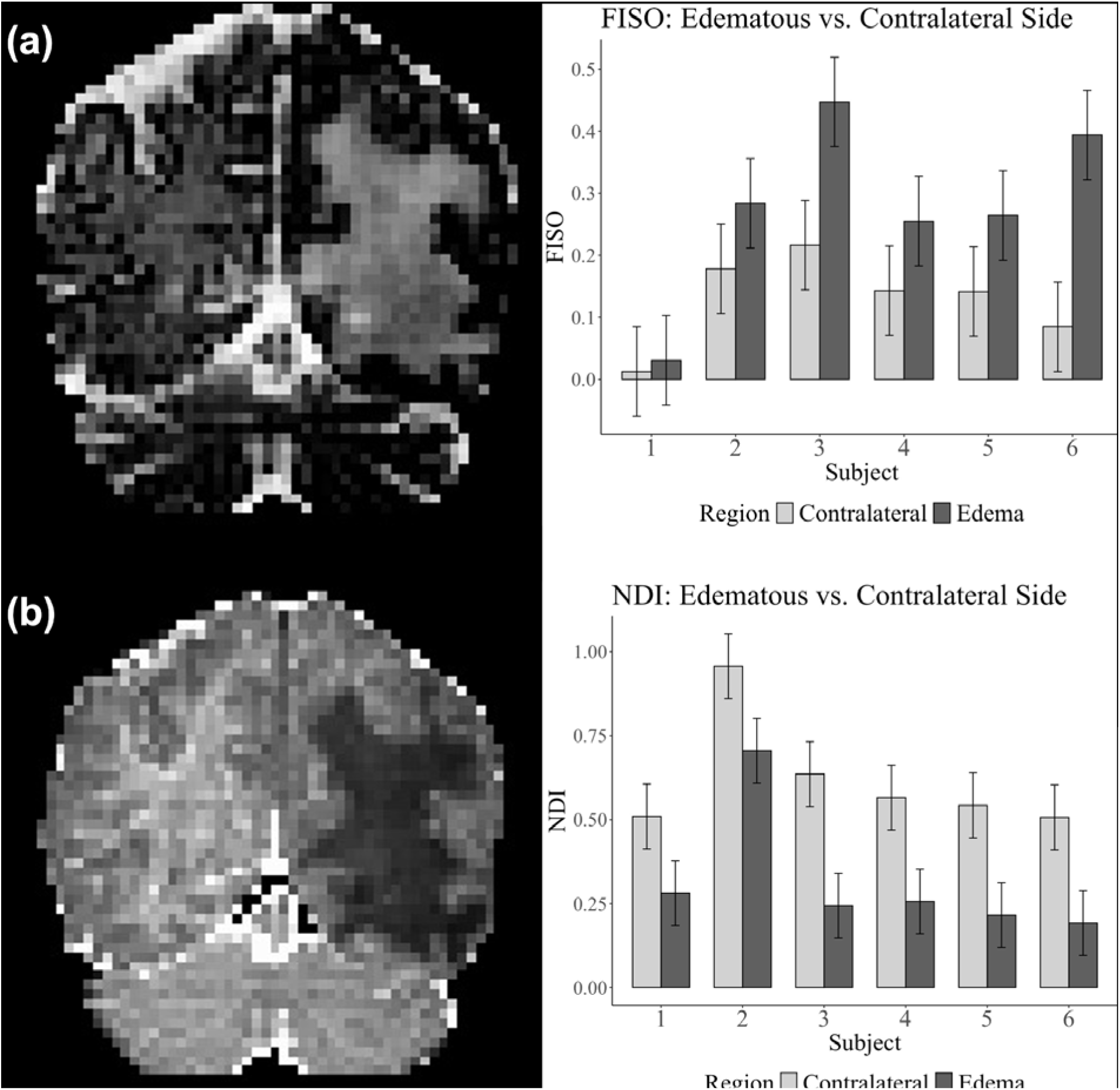
(a) Representative NODDI-derived isotropic volume fraction (FISO) and (b) neurite density index (NDI) maps for a participant with meningioma-induced edema. Plots represent voxel-wise (a) FISO and (b) NDI values in regions of edema versus regions in the contralateral healthy-appearing side of the brain. FISO values were found to be higher in the edematous region, likely due to an increased volume of tumor-induced edema. In contrast, NDI values were found to be lower in edematous regions.

## References

1. Gondar R, Patet G, Schaller K, Meling TR. Meningiomas and Cognitive Impairment after Treatment: A Systematic and Narrative Review. Cancers. 2021;13(8):1846. doi:10.3390/cancers13081846

2. Duffau H. Damaging a few millimeters of the deep white matter tracts during glioma surgery may result in a large-scale brain disconnection. Published online August 11, 2023. doi:10.3171/2023.6.JNS231048

3. Chen Z, Tie Y, Olubiyi O, et al. Corticospinal tract modeling for neurosurgical planning by tracking through regions of peritumoral edema and crossing fibers using two-tensor unscented Kalman filter tractography. Int J Comput Assist Radiol Surg. 2016;11(8):1475–1486. doi:10.1007/s11548-015-1344-5

4. Gong S, Zhang F, Norton I, et al. Free water modeling of peritumoral edema using multi-fiber tractography: Application to tracking the arcuate fasciculus for neurosurgical planning. PLOS ONE. 2018;13(5):e0197056. doi:10.1371/journal.pone.0197056

5. Kaal ECA, Vecht CJ. The management of brain edema in brain tumors: Curr Opin Oncol. 2004;16(6):593–600. doi:10.1097/01.cco.0000142076.52721.b3

6. Lecoeur J, Caruyer E, Elliott M, Brem S, Macyszyn L, Verma R. Addressing the Challenge of Edema in Fiber Tracking.

7. Liao R, Ning L, Chen Z, et al. Performance of unscented Kalman filter tractography in edema: Analysis of the two-tensor model. NeuroImage Clin. 2017;15:819–831. doi:10.1016/j.nicl.2017.06.027

8. Zhang H, Wang Y, Lu T, et al. Differences Between Generalized Q-Sampling Imaging and Diffusion Tensor Imaging in the Preoperative Visualization of the Nerve Fiber Tracts Within Peritumoral Edema in Brain. Neurosurgery. Published online September 19, 2013. doi:10.1227/NEU.0000000000000146

9. Maier SE, Sun Y, Mulkern RV. Diffusion imaging of brain tumors. NMR Biomed. 2010;23(7):849–864. doi:10.1002/nbm.1544

10. Yamasaki F, Kurisu K, Satoh K, et al. Apparent Diffusion Coefficient of Human Brain Tumors at MR Imaging1. Radiology. Published online June 1, 2005. doi:10.1148/radiol.2353031338

11. Hogea C, Davatzikos C, Biros G. Modeling Glioma Growth and Mass Effect in 3D MR Images of the Brain. In: Ayache N, Ourselin S, Maeder A, eds. Medical Image Computing and Computer-Assisted Intervention – MICCAI 2007. Springer; 2007:642–650. doi:10.1007/978-3-540-75757-3_78

12. Basser PJ, Pierpaoli C. Microstructural and physiological features of tissues elucidated by quantitative-diffusion-tensor MRI. J Magn Reson. 2011;213(2):560–570. doi:10.1016/j.jmr.2011.09.022

13. Coelho S, Pozo JM, Jespersen SN, Frangi AF. Optimal Experimental Design for Biophysical Modelling in Multidimensional Diffusion MRI. In: Shen D, Liu T, Peters TM, et al., eds. Medical Image Computing and Computer Assisted Intervention – MICCAI 2019. Lecture Notes in Computer Science. Springer International Publishing; 2019:617–625. doi:10.1007/978-3-030-32248-9_69

14. Jelescu IO, Palombo M, Bagnato F, Schilling KG. Challenges for biophysical modeling of microstructure. J Neurosci Methods. 2020;344:108861. doi:10.1016/j.jneumeth.2020.108861

15. Tropine A, Vucurevic G, Delani P, et al. Contribution of Diffusion Tensor Imaging to Delineation of Gliomas and Glioblastomas. J Magn Reson Imaging. 2004;20:905–912. doi:10.1002/jmri.20217

16. Yen PS, Teo BT, Chiu CH, Chen SC, Chiu TL, Su CF. White matter tract involvement in brain tumors: a diffusion tensor imaging analysis. Surg Neurol. 2009;72(5):464–469. doi:10.1016/J.SURNEU.2009.05.008

17. Qiu A, Alexander AL, Edwards LJ, et al. NODDI-DTI: Estimating Neurite Orientation and Dispersion Parameters from a Diffusion Tensor in Healthy White Matter. Front Neurosci Wwwfrontiersinorg. 2017;11. doi:10.3389/fnins.2017.00720

18. Pasternak O, Sochen N, Gur Y, Intrator N, Assaf Y. Free water elimination and mapping from diffusion MRI. Magn Reson Med. 2009;62(3):717–730. doi:10.1002/mrm.22055

19. Coelho S, Baete SH, Lemberskiy G, et al. Reproducibility of the Standard Model of diffusion in white matter on clinical MRI systems. Published online February 4, 2022. Accessed February 6, 2024. http://arxiv.org/abs/2202.02399

20. Zhang H, Schneider T, Wheeler-Kingshott CA, Alexander DC. NODDI: Practical in vivo neurite orientation dispersion and density imaging of the human brain. NeuroImage. 2012;61(4):1000–1016. 10.1016/j.neuroimage.2012.03.072

21. Wang N, Zhang J, Cofer G, et al. Neurite orientation dispersion and density imaging of mouse brain microstructure. Brain Struct Funct. 2019;224(5):1797–1813. doi:10.1007/s00429-019-01877-x

22. Chong ST, Liu X, Kao HW, et al. Exploring Peritumoral Neural Tracts by Using Neurite Orientation Dispersion and Density Imaging. Front Neurosci. 2021;15. doi:10.3389/fnins.2021.702353

23. Kadota Y, Hirai T, Azuma M, et al. Differentiation between glioblastoma and solitary brain metastasis using neurite orientation dispersion and density imaging. J Neuroradiol. 2020;47(3):197–202. doi:10.1016/J.NEURAD.2018.10.005

24. Li SH, Jiang RF, Zhang J, et al. Application of Neurite Orientation Dispersion and Density Imaging in Assessing Glioma Grades and Cellular Proliferation. World Neurosurg. 2019;131:e247–e254. doi:10.1016/j.wneu.2019.07.121

25. Mao J, Zeng W, Zhang Q, et al. Differentiation between high-grade gliomas and solitary brain metastases: a comparison of five diffusion-weighted MRI models. BMC Med Imaging. 2020;20(1). doi:10.1186/s12880-020-00524-w

26. Masjoodi S, Hashemi H, Oghabian MA, Sharifi G. Differentiation of Edematous, Tumoral and Normal Areas of Brain Using Diffusion Tensor and Neurite Orientation Dispersion and Density Imaging. J Biomed Phys Eng. 2018;8(3):251–260.

27. Maximov II, Tonoyan AS, Pronin IN. Differentiation of glioma malignancy grade using diffusion MRI. Phys Med. 2017;40:24–32. doi:10.1016/j.ejmp.2017.07.002

28. Okita Y, Takano K, Tateishi S, et al. Neurite orientation dispersion and density imaging and diffusion tensor imaging to facilitate distinction between infiltrating tumors and edemas in glioblastoma. Magn Reson Imaging. 2023;100:18–25. doi:10.1016/j.mri.2023.03.001

29. Pieri V, Sanvito F, Riva M, et al. Along-tract statistics of neurite orientation dispersion and density imaging diffusion metrics to enhance MR tractography quantitative analysis in healthy controls and in patients with brain tumors. Hum Brain Mapp. 2021;42(5):1268–1286. doi:10.1002/hbm.25291

30. Wen Q, Kelley DAC, Banerjee S, et al. Clinically feasible NODDI characterization of glioma using multiband EPI at 7 T. NeuroImage Clin. 2015;9:291–299. doi:10.1016/j.nicl.2015.08.017

31. Würtemberger U, Rau A, Reisert M, et al. Differentiation of Perilesional Edema in Glioblastomas and Brain Metastases: Comparison of Diffusion Tensor Imaging, Neurite Orientation Dispersion and Density Imaging and Diffusion Microstructure Imaging. Cancers. 2023;15(1). doi:10.3390/cancers15010129

32. H. J. Scherer, M.D. STRUCTURAL DEVELOPMENT IN GLIOMAS. Am J CANCER. 1938;E XXXIV(3).

33. Cheng-Hong Toh, Alex M-C Wong, Kuo-Chen Wei, Shu-Hang Ng, Ho-Fai Wong, Yung-Liang Wan. Peritumoral Brain Edema in Meningiomas□: Correlation of Radiologic and Pathologic Features. Published online 2011. doi:10.3340/jkns.2011.49.1.26

34. Tournier JD, Smith R, Raffelt D, et al. MRtrix3: A fast, flexible and open software framework for medical image processing and visualisation. NeuroImage. 2019;202:116137. doi:10.1016/j.neuroimage.2019.116137

35. Veraart J, Novikov DS, Christiaens D, Ades-aron B, Sijbers J, Fieremans E. Denoising of diffusion MRI using random matrix theory. NeuroImage. 2016;142:394–406. doi:10.1016/j.neuroimage.2016.08.016

36. Schilling KG, Blaber J, Huo Y, et al. Synthesized b0 for diffusion distortion correction (Synb0-DisCo). Magn Reson Imaging. 2019;64:62–70. doi:10.1016/J.MRI.2019.05.008

37. Jenkinson M, Beckmann CF, Behrens TEJ, Woolrich MW, Smith SM. FSL. NeuroImage. 2012;62(2):782–790. doi:10.1016/j.neuroimage.2011.09.015

38. FSL Diffusion Toolbox Practical. Accessed January 26, 2024. https://fsl.fmrib.ox.ac.uk/fslcourse/2019_Beijing/lectures/FDT/fdt1.html

39. NYU-DiffusionMRI/SMI. Published online June 16, 2025. Accessed June 19, 2025. https://github.com/NYU-DiffusionMRI/SMI

40. Microstructure Imaging Group | Download NODDI matlab toolbox. Accessed January 25, 2024. http://mig.cs.ucl.ac.uk/index.php?n=Download.NODDI

41. Parker C. Not all voxels are created equal: reducing estimation bias in regional NODDI metrics using tissue-weighted means. Chris Parker - Research Blog. November 16, 2021. Accessed November 14, 2024. https://csparker.github.io/research/2021/11/16/Tissue-weighted-mean.html

42. Veale T. tdveale/TissueWeightedMean. Published online March 21, 2024. Accessed November 14, 2024. https://github.com/tdveale/TissueWeightedMean

43. R: The R Project for Statistical Computing. Accessed January 25, 2024. https://www.r-project.org/

44. tckgen — MRtrix 3.0 documentation. Accessed October 26, 2023. https://mrtrix.readthedocs.io/en/dev/reference/commands/tckgen.html

45. imeka/mi-brain. Published online July 6, 2024. Accessed August 2, 2024. https://github.com/imeka/mi-brain

46. Fukutomi H, Glasser MF, Murata K, et al. Diffusion Tensor Model links to Neurite Orientation Dispersion and Density Imaging at high b-value in Cerebral Cortical Gray Matter. Sci Rep. 2019;9(1):12246. doi:10.1038/s41598-019-48671-7

47. Wilkins B, Lee N, Gajawelli N, Law M, Leporé N. Fiber estimation and tractography in diffusion MRI: Development of simulated brain images and comparison of multi-fiber analysis methods at clinical b-values. NeuroImage. 2015;109:341–356. doi:10.1016/j.neuroimage.2014.12.060

48. GCS Ramsay Santé pour l’Enseignement et la Recherche. Evaluation of the Contribution of NODDI Protocol Tractography in Brain Tumor Surgery. clinicaltrials.gov; 2022. Accessed December 31, 2023. https://clinicaltrials.gov/study/NCT05669326

49. Mori S, Crain BJ, Chacko VP, van Zijl PC. Three-dimensional tracking of axonal projections in the brain by magnetic resonance imaging. Ann Neurol. 1999;45(2):265–269. doi:10.1002/1531-8249(199902)45:2<265::aid-ana21>3.0.co;2-3

50. Jeurissen B, Tournier JD, Dhollander T, Connelly A, Sijbers J. Multi-tissue constrained spherical deconvolution for improved analysis of multi-shell diffusion MRI data. NeuroImage. 2014;103:411–426. doi:10.1016/J.NEUROIMAGE.2014.07.061

51. Kamiya K, Hori M, Aoki S. NODDI in clinical research. J Neurosci Methods. 2020;346. doi:10.1016/j.jneumeth.2020.108908

52. Kamagata K, Andica C, Uchida W, et al. Advancements in Diffusion MRI Tractography for Neurosurgery. Invest Radiol. Published online September 15, 2023. doi:10.1097/RLI.0000000000001015

53. Kumpulainen V, Merisaari H, Copeland A, et al. Effect of number of diffusion-encoding directions in diffusion metrics of 5 year olds using tract-based spatial statistical analysis. Eur J Neurosci. 2022;56(6):4843–4868. doi:10.1111/ejn.15785

54. Blaimer M, Choli M, Jakob PM, Griswold MA, Breuer FA. Multiband phase-constrained parallel MRI. Magn Reson Med. 2013;69(4):974–980. doi:10.1002/mrm.24685

55. Deshmane A, Gulani V, Griswold MA, Seiberlich N. Parallel MR imaging. J Magn Reson Imaging. 2012;36(1):55–72. doi:10.1002/jmri.23639

56. Lee DJ, Yoo J, Ye JC. Deep artifact learning for compressed sensing and parallel MRI. Published online 2017. arXiv:1703.01120

57. Palombo M, Ianus A, Guerreri M, et al. SANDI: A compartment-based model for non-invasive apparent soma and neurite imaging by diffusion MRI. NeuroImage. 2020;215:116835. doi:10.1016/J.NEUROIMAGE.2020.116835

58. Jelescu IO, de Skowronski A, Geffroy F, Palombo M, Novikov DS. Neurite Exchange Imaging (NEXI): A minimal model of diffusion in gray matter with inter-compartment water exchange. NeuroImage. 2022;256:119277. doi:10.1016/J.NEUROIMAGE.2022.119277

